# The impact of COVID-19 on community-dwelling people post-stroke and informal caregivers: a qualitative study

**DOI:** 10.1101/2023.07.20.23292901

**Authors:** Teresa Dias, Patrícia Cravo, Joana Santos, Catarina Gomes, Marta Santiago, Carla Mendes Pereira

## Abstract

**Background/ aims:** Little is known about the experience of people post-stroke and their informal caregivers during the COVID-19 pandemic. The aim of this study was to understand the challenges faced by people post-stroke and informal caregivers during the pandemic, as well as the impact on their healthcare support, lifestyle, and self-care behaviors.

**Methods:** A multi-perspective qualitative study was undertaken, with semi-structured interviews being carried out to sixteen participants: eight stroke patients and eight informal caregivers, mostly performed online. Reflexive thematic analysis was used, with data being independently coded and categorized before consolidated into themes and subthemes.

**Findings:** Three themes were derived from the data analysis: i) Perceived impact of COVID-19 pandemic, ii) What helped? - strategies to manage the distress provoked by COVID-19, and iii) The value of rehabilitation and physical activity, with findings highlighting the negative psychological impact of the pandemic. In response to the perceived lack of support and access to health and social services, participants highlighted the use of digital approaches and professional support.

**Conclusions:** Findings suggest the importance of self-management support and/or digital content in order to mitigate the impact of COVID-19. The involvement of peers, family members, friends or others seems to be an important strategy to increase motivation in remote rehabilitation and physical activity.

**Key points:**

- Due to the impact of COVID-19 on people post-stroke and informal caregivers’ daily routines, both highlighted the importance of finding new and alternative ways of communicating, which included the use of digital approaches.
- For some informal caregivers the pandemic was perceived as an opportunity to spend more time with the family and with the person with stroke.
- People post-stroke admit having decreased their levels of physical activity during COVID-19 and increased the value given to rehabilitation and physical activity.
- Involving peers or others, digitally or in-person, seems to be an important strategy when planning physical activity recommendations.

## Introduction

Since the World Health Organization declared the pandemic of Coronavirus 2019 (WHO, 2020), more than 598 million people have been infected with SARS-CoV-2/ COVID-19, and over more than 6 million deaths have been reported around the world (Johns Hopkins University & Medicine, 2022). In Portugal, more than 5 million cases and 24 thousand deaths were registered between March 2020 and August 2022 (DGS, 2022). To mitigate the spread of COVID-19, lockdowns (national and local) and strict social distancing measures have been imposed in many countries, including Portugal (Tanne et al, 2020). Consequently, this unprecedent scenario has reached implications for many aspects of society, including health and economy (Chakraborty and Maity, 2020). Healthcare services were exposed to significant challenges (Mayor, 2020), including the delivery of health care to people post-stroke (Lee et al 2021).

Worldwide, in 2019 stroke remained the second-leading cause of mortality, with 6.55 million deaths from stroke, and the third-leading cause of mortality and disability combined, measured by disability-adjusted life-years (DALYs), with 143 million DAYLS due to stroke (Abbafati et al, 2020; Feigin et al, 2021). Additionally, people post-stroke seem to be more susceptible to COVID-19 (Aggarwal et al 2020), which requires a strict protection. However, the pandemic brought a broader impact on stroke care (Markus Brainin, 2020). World Stroke Organization members reported a significant service reorganization with the reallocation of neurology and stroke beds, including Intensive Care Unit facilities to COVID-19 patients, requiring a shift of stroke units to less optimal accommodations and a redeployment of healthcare professionals (Bersano and Pantoni, 2020; Markus and Brainin, 2020). A delayed admission to stroke units and less specialized stroke services were also reported (Alonso De Leciñana et al, 2021; Markus and Brainin, 2020; Markus and Martins, 2021). Many healthcare systems have reduced or suspended “non-urgent” care and discharged patients earlier than usual (Bettger et al 2020). Changes in post-acute care patterns were observed with more patients being discharged to home (Srivastava et al 2021) and fewer discharged to inpatient rehabilitation facilities (Thau et al 2021). These changes also affected the role and care demands from the perspective of caregivers, with difficulties related to limited communication with health professionals, decrease of adequate training on procedures and increase of their workload (Lee et al 2021). Similarly, outpatient clinics and rehabilitation services have reduced or stopped patient services (Alonso De Leciñana et al 2021). Although several services increasingly started to use telerehabilitation platforms, this type of assistance may present barriers with difficulties with technology use for the generally old population of stroke patients (Bersano et al 2020).

Even before the pandemic, it is well-documented in literature that people post-stroke and their informal caregivers, usually family members or friends, often feel abandoned and marginalized by healthcare services (Pindus et al 2018). Stroke caregivers witnessed a crushing care burden, both physical (e.g., fatigue) and psychological (e.g., anxiety and depression) (Denham et al 2020; Hodson et al 2020; Luker et al 2017), which increases with longer hours of care (Gbiri et al 2015). Moreover, in Portugal there has been a traditional reliance on the family as the first line of informal care for dependent people (Barbosa et al 2020), with home-caregiving and stroke-related consequences being more disruptive among Portuguese family caregivers, compared with other countries of Northern Europe (Lurbe-Puerto, Leandro Baumann, 2012; Pereira Rebelo Botelho, 2011).

Recent studies reported that the COVID-19 pandemic causes a reduction in overall quality of life, an increase in anxiety, fear and feelings of social isolation among the general public (Mukhtar, 2020; Saltzman et al, 2020). Similarly, people post-stroke seem to report increased stress, anxiety and decreased levels of health-related quality of life during the pandemic (Almhdawi et al, 2021; Singh et al, 2021). Moreover, chronic patients, including people post-stroke, faced difficulties in accessing healthcare and lived a sedentary lifestyle (Singh et al 2021). However, the literature is not clear, with findings revealing that in the context of inpatient rehabilitation, patients may feel more secure due to the variety of prevention measures, and taking to higher levels of health-related quality of life during the pandemic than before (Zhao et al 2021). Furthermore, the extra time spent at home by patients and their family members during the pandemic also showed some positive effects, such as a perceived benefit of spending “more quality time with family” (Fama et al 2021). However, an increase in video chats with family and friends has shown to be negatively correlated with perceptions of friendships, which may be explained by some frustration with technology and in comparison with in-person interactions (Fama et al 2021).

In addition, the COVID-19 pandemic provoked significant changes in the routines of family caregivers of community-dwelling people post-stroke. Recent studies show that caregivers’ burdens seem to be exacerbated by the pandemic (Lee et al 2021; Sutter-Leve et al 2021). One study with caregivers of individuals with a newly-acquired stroke admitted to inpatient rehabilitation unit, in the United States, reported a COVID-19 impact on competency of care, with participants feeling unprepared for discharge and reporting communication challenges (Sutter-Leve et al 2021). Another study conducted in China (Lee et al 2021), reported that stroke caregivers during the pandemic had to deal with care service adversities, additional caregiving workload and strain, had threatened relationship between the caregiver and the person post-stroke, threats to caregivers’ physical and psychological well-being, and needs for continuing caregiving roles.

To our knowledge, studies have been focused on people post-stroke or family caregivers separately. However, the global impact of COVID-19 pandemic measures and the importance of family support after a stroke highlights the need for a dyadic perspective in understanding the impact within the family. In addition, there are no studies conducted in a Portuguese setting. Therefore, this study aimed to explore and understand the challenges faced by Portuguese people post-stroke and informal caregivers during the pandemic, as well as the impact on their healthcare support, lifestyle, and self-care behaviors. This study intended to answer the research question: How has the COVID-19 pandemic affected both community-dwelling people post-stroke and informal caregivers?

## Methods

### Study design

A multi-perspective qualitative study was adopted, following an constructivist-interpretivist paradigm (Denzin Lincoln, 2018). Due to the nature of data being sought, a reflexive thematic analysis approach was used through in-depth semi-structured interviews (Braun Clarke, 2021), in order to allow flexibility in response to the participant being interviewed and the interview context. In a pandemic context, a qualitative approach may provide insight into people’s lived experiences of disease and their coping strategies, which complements epidemiological data from quantitative research (Vindrola-Padros et al 2020). Moreover, a digital qualitative approach was undertaken in order to increase the opportunity for people to fully participate without being exposed to COVID-19. The standards for reporting qualitative research (SRQR) were followed for this paper (O’Brien et al 2014).

By adopting a multi-perspective approach (Kendall et al 2010), this research allowed an in-depth understanding of the impact of COVID-19 pandemic on the everyday life of both people post-stroke and informal caregivers. Previous studies from a family or couples’ perspectives underline the impact of stroke on both the person who has had a stroke and their family and the role of interdependence between them, indicating that stroke should not be seen as an individual phenomenon (Pereira, Greenwood Jones, 2020; Theadom, Rutherford, Kent McPherson, 2018).

Topic interview’s guides focused on: 1) the challenges faced during the COVID-19 pandemic and their impact on participants’ access to healthcare, lifestyle, mental and self-care behaviors (e.g., exercise); 2) the participants’ unmet needs during the COVID-19 pandemic, and 3) the strategies used by participants to minimize the impact of the pandemic, recommendations to healthcare services/ professionals and key lessons learnt from the situation. The guides were developed by the primary researcher (CP) in consultation with the wider research team and were pilot tested in order to assure the adequacy of the questions to the research topic, with the final main topics adapted for people post-stroke and informal caregivers (Malmqvist et al., 2019).

Ethical approval was obtained from the Ethics Committee of the Health School of the Polytechnique Institute of Setubal (66/AFC/2020). An information sheet was given to each participant and details about the purpose, nature and procedures of the study were explained. Participants who agreed to take part of the study signed then an online consent form (Salmons, 2016). Confidentiality and anonymity of the participants was guaranteed and all the data that could lead to a possible identification was kept confidential.

### Participants

Participants were recruited from social media. The study was promoted digitally using Facebook. Recruiting via traditional methods may have been difficult in the context of lockdowns and disruption due to the COVID-19 pandemic. Considering the visibility and Facebook users in Portugal (around 8 million users), recruitment posts were published in the Facebook accounts of two national associations of patients and informal caregivers (PT.AVC-União de Sobreviventes, Familiares e amigos, literally “PT.Stroke-Union of Survivors, Family and Friends”, and ANCI-Associação Nacional de Cuidadores Informais, literally “National Association of Informal Caregivers”), and disseminated by the social media of the Health School of the Polytechnique Institute of Setubal.

For people post-stroke, the following inclusion criteria were used: 1) diagnosis of stroke from clinical observation or brain imaging; 2) age over 18 years old; 3) living at home, with internet access and a mobile device allowing access to videoconference calls; and 4) ability to communicate in Portuguese. Exclusion criteria were severe aphasia, dementia and illiteracy. Informal caregivers, defined as the person who was supporting the person post-stroke at home, without payment for that service (Diário da República, 2020), were included if: 1) they took care of a person who has had a stroke; 2) were over 18 years old; and 3) had internet access and a mobile device allowing access to videoconference calls. They were excluded if the person who they took care of was hospitalized or in inpatient facilities.

### Data collection

Eligible participants who expressed their willingness to participate by submitting an online consent form were invited to take part of the study. An initial contact was made by email, sending detailed information about the study and proposing a contact by phone to explain the purpose, nature and procedures of the study. During or after the phone call, depending on the participants’ preferences and convenience, the participants who agreed to take part of the study signed then a consent form “click-if-you-agree” type, sent by email, filled a socio-demographic form and a version adapted to the Portuguese language of the Stroke Impact Scale (SIS 3.0) (Teodoro Pereira, 2009). Then, a convenient date, place and time for interview was scheduled with participants. The setting for interviews was chosen according to the participants’ preferences. Interviews carried out mostly online using the Zoom colibri digital platform and were conducted by two female members of the research team (MS, CG), who were specifically trained for this study. One interview was carried out in person and took place at an outpatient clinic. During the online interview, participants were recommended to be at home in a quiet room. The researcher’s camera was kept on, but the microphone was turned off whenever the participant was speaking, in order to avoid interference. No internet connection issues were identified, and a quiet non-disturbed environment was respected during the process.

Additionally, for participants in a close and dyadic relationship (person post-stroke and informal caregiver), separate interviews were planned to protect participants’ privacy and minimize responses perceived as acceptable to the partner (Kendall et al 2010). Exception was considered during one interview with the presence of the husband and granddaughter, without interfering, aiming to support the participant with the use of the digital platform.

Participants were recruited over four months. No one refused to participate in the study. Data were collected between December 2020 and June 2021. Interviews lasted between 15 and 60 minutes and were audio recorded, which allowed preservation of the raw data for review at a later time, and additional notes were taken during interviews.

### Data analysis

All interviews were transcribed verbatim immediately. Data obtained from people post-stroke and caregivers were coded separately followed by an analysis of similarities and differences between the datasets. The two datasets were merged when similar themes were identified. Thematic analysis offered the flexibility required to adopt an initial eagle-eyed approach in identifying general themes, whilst retaining individual contributions of participants from different groups (Braun Clarke, 2006). Therefore, it allowed preserving the context of individual participants and group journeys, as well as looking for patterns threaded through the two different group’s sets.

In these separate processes, two researchers independently familiarized themselves with each dataset and inductively assigned codes line by line (PC, JS). These initial steps of coding interviews from the each group set were followed by the sequence phases in searching themes and sub-themes (Braun Clarke, 2006), which were discussed in regular meetings and crosschecked by other research team members (CP, TD).

The study was iterative, in the sense that data collection and analysis were carried out in parallel and emerging findings shaped the data collection process. An initial analysis took place immediately after the first four interviews, which helped identify the need of gathering insight from the perspective of people who suffered a stroke during the COVID-19 pandemic. Initial codes were mainly focused on the impact of the COVID-19 pandemic of people in a chronic stage after stroke. Thus, efforts were made to recruit participants in sub-acute phase of recovery after stroke, in order to achieve thematic saturation of data, which was analyzed during the debriefing meetings carried out through the analysis process.

### Methods to assure trustworthiness, rigor and credibility

Criteria for assessing research quality were considered according to the philosophical stance of the study (Guba Lincoln, 2005). Reflexivity was carried out as an active acknowledgement and explicit recognition by the researchers about how their position and presence may affect the research process (Berger, 2015). This was crucial to acquire the point-of-view of the participants and let them speak for themselves, while allowing the researchers to continuously understand their own perspective (Berger, 2015; Calman et al 2013). Peer debriefing was used during the analysis process with regular debriefing sessions between the researcher team. Also, there was no previous contact or relationship between interviewers and the participants of the study. The reasons for undertaking the study were explored during the explanation of the study aims and procedures before participants consent for participating in the study. The research team was composed of Portuguese female citizens, with background in Physical Therapy. The participants knew the interviewers/facilitators regarding their gender, reasons for doing the study, area of expertise and research interests (physical therapy, neurology and stroke rehabilitation).

## Findings

### Participant Characteristics

A total of sixteen participants were included in the study. From these, eight were people who had experienced a stroke and eight were informal caregivers. Details about the socio-demographic characteristics of the participants are provided in Tables 1-3.

**Table 1.** Characteristics of the study populations included in the study.

|  | N | Age in years<br>mean (SD) | Gender<br>female | Educational level<br>N (%) | Professional status<br>N (%) |
| --- | --- | --- | --- | --- | --- |
| **Participants (total)** | 16 | 52.56 (19.13) | 12 | Primary= 2 (12.5%)<br>Secondary= 7 (43.75%)<br>High school= 7 (43.75%) | Retired= 5 (31.25%)<br>Unemployed= 3 (18.75%)<br>Employed n=8 (50%) |
| **People post-stroke** | 8 | 55.12 (22.75) | 7 | Primary= 1 (12.5%)<br>Secondary= 5 (62.5%)<br>High school= 2 (25%) | Retired= 4 (50%)<br>Unemployed= 3 (37.5%)<br>Employed n=1(12.5%) |
| **Carers** | 8 | 50 (14.85) | 5 | Primary= 1 (12.5%)<br>Secondary= 2 (25%)<br>High school= 5 (62.5%) | Retired= 1 (12,5%)<br>Employed n=7 (87.5%) |

Legend: N- number; SD- Standard Deviation

**Table 2.**
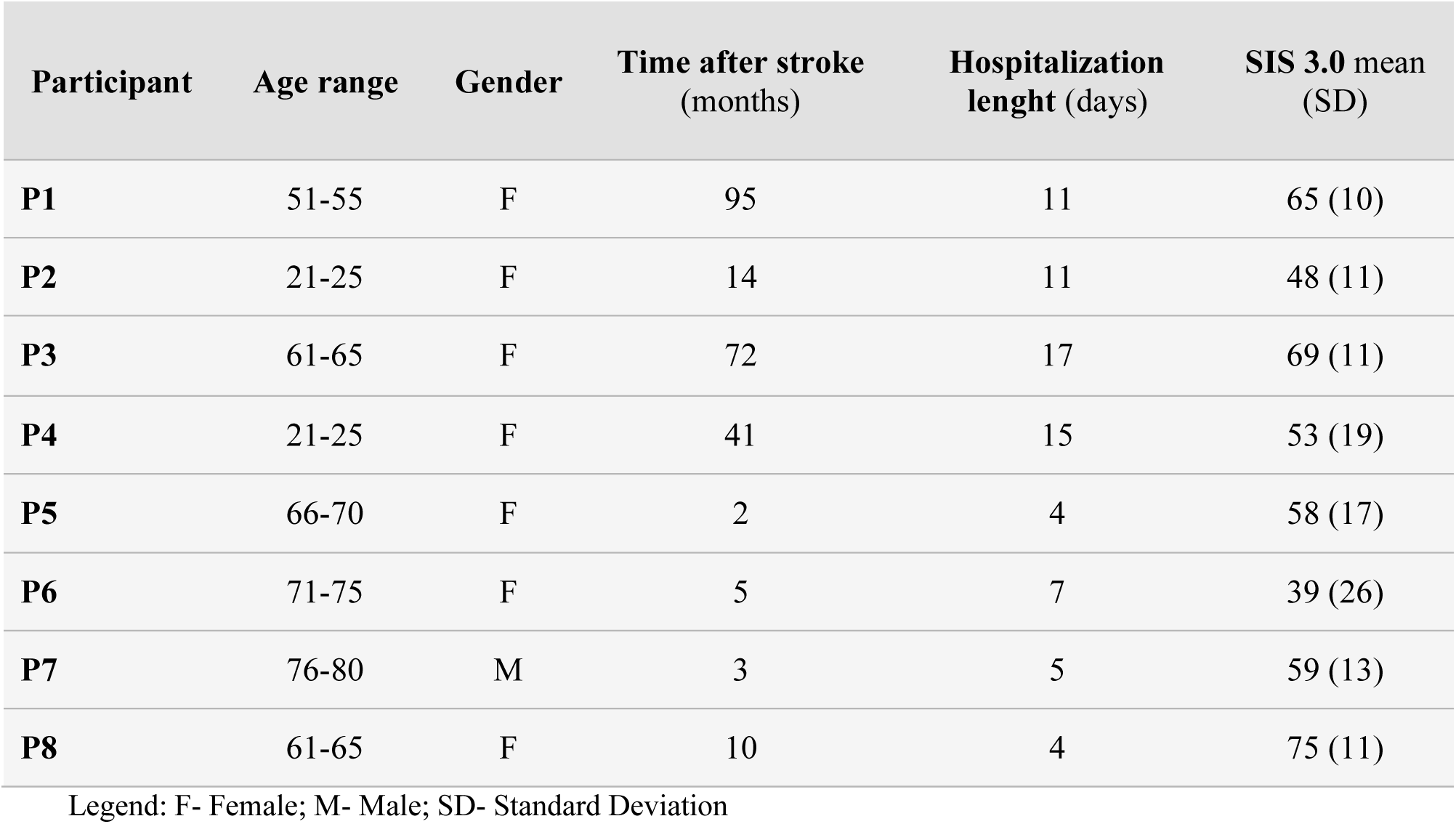
Characteristics of people post-stroke (n=8).

**Table 3.** Characteristics of informal caregivers (n=8).

| Participant | Age range | Gender | Relationship with the PPS | Living situation | Time as caregiver (months) | Previous experience of caring | Time spent on care (hours per day) |
| --- | --- | --- | --- | --- | --- | --- | --- |
| **P9** | 21-25 | F | Child | Living with the PPS | 9 | Yes | 5 |
| **P10** | 46-50 | F | Relative | -- | 11 | Yes | 8 |
| **P11** | 41-45 | M | Spouse | Living with the PPS | 17 | No | 12 |
| **P12** | 56-60 | F | Spouse | Living with the PPS | 2.5 | No | 3 |
| **P13** | 71-75 | M | Spouse | Living with the PPS | 9 | No | 2 |
| **P14** | 41-45 | M | Child | Living with the PPS | 3 | Yes | 2 |
| **P15** | 66-70 | F | Spouse | Living with the PPS | 2 | Yes | 12 |
| **P16** | 41-45 | F | Child | Living with the PPS | 5 | No | 12 |

Legend: F- Female; M- Male; PPS- Person Post-Stroke

Of the post-stroke people (mean age: 55.1±22.75) 87.5% were female (n=7) and 56% were retired (n=5). The average time after stroke was 30.25 months (± 35.66) with the participants having diverse degrees of impairment and health-related quality of life after stroke assessed by SIS 3.0 (mean 58.25 ±11.62) (Duncan et al 2003). The hospitalization length varied between 4 and 17 days (mean days: 9.25±5.04) with the average time in hospital being higher before the pandemic.

Of all the informal caregivers (mean age: 50±14.85) 62.5% were female (n=5) and 50% were relatives of people post-stroke who participated in the study (n=4; 2 were spouses and 2 were children). Among them, 87.5% were employed, had been caring for 7.25 months on average (±5.3), and 62.5% received additional support (n=5).

### Interview findings

Three major themes derived from the data analysis: i) *Perceived impact of COVID-19 pandemic*, ii) *What helped?-strategies to manage the distress provoked by COVID-19,* and iii) *The value of rehabilitation and physical activity.* Each of the three major themes was connected to sub-themes, as detailed in table 4 and described below.

**Table 4.** Themes and sub-themes.

| Themes | Sub-themes |
| --- | --- |
| Perceived impact of the COVID-19 pandemic | Changed routines, added uncertainties |
|  | Lack of access and support from social and health services |
| What helped? - strategies to manage distress provoked by COVID-19 | Digital solutions and new ways of communicating |
|  | Empathy from health professionals |
| The value of rehabilitation and physical activity | Perceived importance of rehabilitation and physical activity |
|  | Barriers to physical activity |

In the theme “perceived impact of COVID-19 pandemic” two key issues were explored by participants, namely, the impact of COVID-19 on both people post-stroke and informal caregivers’ daily routines and the perceived lack of support and access to social and health services due to service reorganizations.

The theme “what helped? - strategies to manage the distress provoked by COVID-19” highlights the importance of finding new and alternative solutions to the challenges added by the pandemic situation, which included the use of digital approaches. For both people post-stroke and informal caregivers, the compassion and empathy from health professionals was also perceived as particularly valuable during these difficult times.

During this pandemic period, people post-stroke and caregivers’ viewpoints about the rehabilitation and physical activity seemed to change, with the theme “the value of rehabilitation and physical activity” suggesting an increased value given to rehabilitation and physical activity. Despite this, for some people significant barriers were difficult to overcome, emphasizing the importance of professional support.

### Perceived impact of the COVID-19 pandemic

#### Changed routines and added uncertainties

From the participants’ perspective, the pandemic provoked significant changes in their daily routines and in how they managed problems and challenges related to stroke. Their lifestyle was affected, with almost all participants reporting that they were feeling quite isolated due to the restrictions imposed by the government. Some participants mentioned going outside only to go to the supermarket and have reduced most outdoor activities. Moreover, the atmosphere of uncertainty and fear related to COVID-19 was perceived as an added problem. Participants reported that this pandemic situation aggravated their previous difficulties in dealing with the overwhelming consequences of the stroke. These added uncertainties, fears and social isolation seemed to lead to significant psychological consequences, particularly in people post-stroke, with most participants reporting feeling down for not maintaining physical contact with other people, either close friends or from rehabilitation settings. Fear of contracting COVID-19 and feelings of stress and anxiety related to the uncertainties and lack of control over the situation were also reported by participants.

> *“At least for me, (the pandemic) changed everything (gets emotional). Psychologically… from the moment that you can no longer see people. You no longer can have contact with people, you have to stay at home, and you don’t know the future.”* (Participant 1, person post-stroke)

The majority of informal caregivers highlighted the impact of the increased time dedicated to care for their relatives, which added difficulties in their time management. From the informal caregivers’ perspective, the uncertainties and lack of social responses had a negative impact on their daily care routines, with some having to reduce their work time or even stop working in order to take care of the person post-stroke.

> *“I don’t have much time for myself, Ím more tired, of course (…). I don’t know how it’s going to be, if I’m going to be able to keep reconciling the care I give and keep working.”* (Participant 11, informal caregiver)

Although most caregivers perceived some level of helplessness and feeling alone in their role as informal caregivers, some looked at the situation as an opportunity to spend more time with the person post-stroke. Flexible work practices and working from home to reduce the spread of the disease were also perceived as increasing flexibility to take care of their relative or to have extra support from other family members, as mentioned by a caregiver, whose daughter helped during this period:

> “*My daughter had online lectures. As she was at home, she took the opportunity to help her grandmother.*” (Participant 14, informal caregiver)
>
> “*In a way, it even made it easier because now I’m working from home. If I was at the office, I wouldn’t have the possibility of helping my mother so much (…). At home I already have more… more time for her and for everything I must do.*” (Participant 16, informal caregiver)

### Lack of access and support from health and social services

Almost all participants reported negative experiences related to the health system during the pandemic. Early discharges due to service reorganizations or to positive cases for COVID-19 in the service/ ward or stroke unit, and lack of information and education at the moment of hospital discharge were commonly reported by both people post-stroke and informal caregivers. Moreover, difficulties in communication within healthcare teams or with the patient and/or informal caregiver and the cessation of “non-urgent” services, such as consultations, speech and language therapy or physical therapy seemed to lead to feelings of abandonment. Some participants also perceived these disruptions as having a negative impact on recovery or even worsening their condition.

> *“I’m sure that under normal circumstances, he would certainly have other type of treatment and would obviously be much better.”* (Participant 10, informal caregiver)
>
> *“They* (health professionals) *didn’t even say overnight that we were going out. It was all in a hurry. We left… and that’s it, with no tips or anything. It was all very sudden.”* (Participant 5, person post-stroke)
>
> *“They* (health professionals) *just said: “come here and get her, she will be discharged tomorrow”* (…) *We didn’t even know how she was, how was her mobility… nothing.”* (Participant 16, informal caregiver)

Furthermore, since the pandemic some participants started avoiding public health services, which was described by some as related to their experience during the hospitalization after the stroke. However, for others this avoidance appeared to be justified by the idea created through television news and social media of chaos in public hospitals.

> *“Everything inside the hospital made me a lot of confusion. So, the way things were, I didn’t go back there.”* (Participant 6, person post-stroke)
>
> *“We asked for help, but not from the national health service because we were in a pandemic… you know how things were, we all watched it on the news… with all the ambulances in front of the hospitals* (Participant 9, informal caregiver)

The lack of governmental solutions to overcome the social needs provoked by restrictions imposed in response to the COVID-19 pandemic were also reported by participants. Most of the social services were closed during lockdowns, which significantly reduced the support usually given to both people post-stroke and informal caregivers. Some participants highlighted the absence of social/governmental support, with the help that they used to receive from public services being suspended. This situation seemed to add more uncertainty, anxiety, irritation, frustration and helplessness feelings in informal caregivers about how to manage all the needs of their relatives.

> *“If the day centers will be closed again, he will no longer have access to this service. And then, how is going to be?”* (Participant 10, informal caregiver)
>
> *“Support was given to parents of children up to 12 years old. Why is this type of support not given to informal caregivers?”* (Participant 16, informal caregiver)
>
> *“Therapy sessions are going to be suspended again because hospitals are closed (…). And this is obviously unknown. We are very scared about it.”* (Participant 15, informal caregiver)

### What helped? - strategies to manage the distress provoked by COVID-19

#### Digital solutions and new ways of communicating

The lack of social support forced some participants to look for new solutions and find new ways of taking care of their relatives. Despite the recognized effort from informal caregivers in their care role and the feeling of a natural responsibility, it seemed to be perceived as not being enough or even possible to continue providing care on a continuously and daily basis. Finding help from other family members, looking for inpatient institutions or hiring a formal caregiver were some of the solutions found by families in order to manage the overwhelming caregiver role and uncertainties surrounding the pandemic situation.

> *“My daughter has been helping a lot… she and my son-in-law. I don’t know what would be of my wife alone with me.”* (Participant 3, person post-stroke)
>
> *“I had to institutionalize him because my holidays ran out. And that’s why I had… I didn’t see any other solution if not to leave him in the institution. I did everything I could before making this decision. I used all my vacation days during lockdown.”* (Participant 10, informal caregiver)

As reported above by participant 10, not all the decisions appeared easy to make, which seemed to be related to the perceived commitment to the family values.

Digital solutions allowed greater accessibility and provided different ways of communication. On one hand, they were perceived as useful tools to access information in order to learn how to carry out daily tasks at home. For some participants, internet was an important source of knowledge, helping them answer some doubts and becoming competent on their caregiving role, which appeared to counteract the lack of support during discharge from hospital. On the other hand, finding a virtual way of “connecting” with others was perceived as minimizing the risk of contagion and internet was perceived as having a valuable role in entertainment.

> *“For example, two weeks ago I went to IKEA and had a video call with my mom so she could choose what she wanted.”* (Participant 9, informal caregiver)
>
> *“It’s better to entertain ourselves with the internet, with a book and watching a good moving… things like that help to forget about the pandemic.”* (Participant 1, person post-stroke)
>
> *“I was doing what I could. I went to the internet, and I looked for how and what were the best ways to do the personal hygiene, such as, bathing.”* (Participant 16, informal caregiver)
>
> *“We as a family and myself ended up creating a movement that I called “CofiqueARir”, to help my cousin and also my mother who was confined at home*.
>
> *Every day we published activities, with the volunteering of many people. There were neurocognitive activities, yoga sessions, theater… physical exercise. It was what I call a, laugh therapy… Everything online.” (*Participant 10, informal caregiver)

Finding these new and alternative ways of communicating seemed to help minimize feelings of isolation or abandonment. Being virtually with the person post-stroke and others, encouraging them, receiving and giving emotional support appeared to be helpful strategies to reduce the emotional burden during the pandemic.

### Empathy from health professionals

From the perspective of both people post-stroke and informal caregivers, the support given by health professionals was crucial to reduce the participants’ distress particularly during the COVID-19 pandemic. The health professionals’ empathy and compassion were highlighted by participants who felt isolated, with their contact from family and loved ones being cut off. Perceiving that health professionals were sensitive to their thoughts and worries appeared to be of a huge importance for both people post-stroke and informal caregivers in such a difficult moment.

> *“I insisted so much that they (health professionals) ended transferring him to another ward so I could come in and visit him.” (*Participant 13, informal caregiver)
>
> *“It was a head nurse who provided me everything I needed, and I am very grateful to her.”* (Participant 5, person post-stroke)
>
> *“In the pandemic situation, when the visits were canceled, it was a little difficult because what was I left with? Three days maybe without seeing her (mother). But then they (health professional) made the exception and from then on I stayed there all day with her in intensive care.”* (Participant 9, informal caregiver)

When possible, informal caregivers also perceived that the time and training provided by health professionals before discharge was essential, helping them to feel competent to assume their new roles at home. This was recognized as valuable by participants who experienced stroke before the pandemic. However, before and during the pandemic there were participants, both people post-stroke and caregivers, who felt that it would be important to receive guidance and support from health professionals in their transition to home.

> *“I had a lot of help from medical staff, nurses and assistants, hm, which allowed me to gain some skills, especially in terms of hygiene, positioning. At least I was able to acquire some skills.”* (Participant 11, informal caregiver)
>
> *“At least, they* (health professionals) *should gave some tips one or two days before* (discharge)*… what could help us during integration at house. But there was nothing.”* (Participant 4, person-post stroke).

### The value of rehabilitation and physical activity

#### Perceived importance of rehabilitation and physical activity

Both people post-stroke and informal caregivers emphasized the value of rehabilitation in the maintenance and improvement of the clinical condition. They both perceived that the COVID-19 pandemic had a negative impact on the recovery after the stroke, with some participants expressing their concerns about the increased dependence of the person post-stroke during this extended period of service suspension.

> “*For people who… have some motor disability, they need to do physiotherapy (…) for me and for everyone it got worsen during the pandemic because I couldn’t do what I needed to do to improve.”* (Participant 1, person post-stroke)
>
> “*He was without speech therapy for three months, which was critical.*” (Participant 10, informal caregiver)

Having their rehabilitation services suspended seemed to increase the perceived value of rehabilitation in the recovery, which made some participants seek alternative ways of maintaining some level of physical activity. Different solutions, such as opting for outdoor spaces, individual activities (e.g. a walk), following exercise videos from the internet or a home exercise program, were perceived as safe ways to engage in physical activity.

> *“I used to do physical therapy and go to the gym. Since the pandemic, I stopped doing both things. I started doing exercise by myself.”* (Participant 2, person post-stroke)
>
> *“We live in the countryside… and that’s it… during lockdowns I started doing walks around there”* (Participant 4, person post-stroke)
>
> *“I stopped physiotherapy, but I’m doing walks. Only, if it’s raining, I don’t go (…) I choose a place where no one passes. It’s in a public park and if I find someone I use my mask.”* (Participant 1, person post-stroke)

Despite the restrictions imposed during the pandemic situation, receiving some sort of guidance from health professionals seemed to be perceived as important to maintain their focus and motivation for recovery after a stroke.

> *“I have been doing exercise by myself* (…) *but, I didn’t know if I was doing it correctly because I could be doing something wrong… However, in the meantime I also received a plan to do physical therapy at home.”* (Participant 2, person post-stroke)

#### Barriers to physical activity

The majority of people post-stroke reported a reduction in physical activity and increase of sedentary behaviors. Some participants identified the lack of motivation and self-efficacy as a barrier to carry out physical activity alone. For them, doing it with the guidance and feedback from a professional was highlighted as important. Not being able to have their support and interaction with other patients, such as, in exercise groups, was perceived as disheartening, resulting in an increased sedentary lifestyle.

> “*It’s very complicated… I have a dog and I try to go for a walk with her to keep myself active. But it’s very complicated because I lose the motivation to do things (…) the conditions are not the same, we cannot have contact with people, it’s not the same.”* (Participant 2, person post-stroke)
>
> *“Since the pandemic came, I have been here at home and I don’t do anything…I don’t want to. I only go for a walk when the weather is good.*” (Participant 3, person post-stroke)

Moreover, for some participants the benefits of rehabilitation and physical activity appeared not to surpass the fear of infection by Sars-cov2, preferring not to return to previous activities.

*“I used to do physical therapy every day. Then, I stopped doing it. Now, they already called me several times to go back and explained to me the new*

> *rules, but I won’t go back until, until the pandemic ends.”* (Participant 1, person post-stroke)

## Discussion

The current COVID-19 pandemic has had multiple and profound implications in healthcare systems worldwide, in particular for people post-stroke and their informal/ family caregivers. By undertaking a multi-perspective design, this study brings new insights about the meaning and impact of COVID-19 upon people post-stroke and their informal/ family caregivers.

Findings demonstrated that the atmosphere of uncertainty, lack of control and fear affected both people post-stroke and informal caregivers, with previous difficulties in dealing with the burden of stroke being aggravated with the pandemic. Their lifestyle was changed due to the restriction imposed, which made them feel quite isolated. However, giving support to each other appeared to be helpful to reduce the emotional burden. Finding new and alternative ways of communicating at home and during hospitalization were reported by both as important strategies to reduce feelings of abandonment. These feelings represent difficulties previously reported in literature (Pindus et al 2018) and aggravated by the pandemic (Almhdawi et al 2021; Singh et al 2021), for which participants looked for solutions by themselves. Despite findings from the National Statistics Institute showing that only around 50% of the population living in Portugal between 16 and 74 years old have basic or above basic digital skills (INE, 2019), the participants from this study highlighted the role of digital solutions to overcome their needs. These findings are in line with the trend of growth in the use of information and communication technologies (ICT) in the daily life of people post-stroke (Gustavsson et al 2018). The current pandemic has reinforced the importance of both people post-stroke and informal caregivers being able to make informed decisions, with digital skills demonstrating benefits in this study. ICT facilitates health decisions, when individuals can autonomously search for information according to their preferences, choose forms (textual, audio or video) and channels of communication, learn and share knowledge with peers and/or professionals (Du et al 2016; WCPT/INPTRA, 2019). ICT contributes to increase health literacy, which enhances the ability to make the decisions necessary for autonomous health management (Costa et al 2019; De Wit et al 2017). Even before the pandemic, the demand and the opportunity to create a national strategy for health literacy in Portugal was recognized along with the life course integrated in the modernization program of the National Health Service entitled “SNS +Proximidade” (Costa et al 2019). A study conducted in Portugal shows that 61% of the sample had ill-health or inadequate level of health literacy (Pedro et al 2016), which emphasizes the importance of addressing these educational needs in the future.

Furthermore, this study has provided different perspectives about the impact of the COVID-19 pandemic in informal caregivers. Although some informal caregivers reported the increased time needed to care for their family members and reduced self-care time (as also reported in previous studies, e.g. Lee et al, 2021; Sutter-Leve et al, 2021), others reported feeling differently. For these participants, the pandemic was perceived as an opportunity to spend more time with their family and receive extra help from other family members, working and studying from home. Similar findings were reported from the perspective of people post-stroke by Fama et al (2021), but not from the perspective of informal caregivers as a positive change. In their belief, this has happened due to flexible work practices, including the possibility to work from home. These findings may be explained by a strong commitment to the family, culturally justified and socially accepted. Other Portuguese findings also show the devotion in providing care to the dependent relative (Pereira and Rebelo Botelho, 2011) and the importance of mutual support within the family (Lurbe-Puerto et al, 2012; Pereira et al, 2020).

In the present study participants reported negative experiences about the healthcare system during the pandemic with earlier discharges, communication difficulties between patients/caregivers and healthcare team, cessation of “non-urgent” services, lack of information and follow-up support leading to feelings of abandonment. These changes compromised also caregivers’ ability to provide safe and effective care to their family members, which was also reported in previous studies (Lee et al, 2021; Sutter-Leve et al, 2021; Zafra-Tanaka et al, 2022). It is well documented in the literature that people post-stroke and their informal caregivers value the support of health services, including learning about the condition and how to deal with it over time in order to take informed decisions about their needs (Chen et al, 2021; Lee et al, 2021; Pereira et al, 2020; Pindus et al, 2018; Theadom et al, 2018). In addition, participants also experienced a lack of social and governmental solutions, expressing their concerns about the reduction of the support usually given (e.g., day care services, formal caregiving services), the need for caregivers to provide full-time care and the consequences of not working. This scenario was also identified around the world, particularly in China, USA, India, and Italy (Budnick et al, 2021; Lee et al, 2021; Singh et al, 2021; Sutter-Leve et al, 2021). In Portugal this scenario occurred under previous circumstances identified as more disruptive and with a higher percentage of co-residential caregivers with depressive symptoms compared to other European countries (Barbosa et al, 2020; Lurbe-Puerto et al, 2012), which may be aggravated during the pandemic.

In this study, both people post-stroke and their informal caregivers highlighted the value and role of rehabilitation in the improvement of the clinical condition. Some participants considered rehabilitation disruption as having a negative impact on their functional recovery and even worsening their condition, as also reported in previous studies (Lee et al, 2021; Pelicioni et al, 2020; Sutter-Leve et al, 2021). In order to overcome these impacts, digital interventions have been proposed to show promising results, although they need attention for neurological patients for safety reasons (Pelicioni et al, 2020). The pandemic introduced numerous changes in the stroke patient’s multidisciplinary rehabilitation, like: hospital visits suspended, replaced with virtual contact using iPad; use of facemasks and visors; community clinic spaces closed; local lockdowns and public health restrictions closing local community resources; virtual support groups (Lucas et al, 2021). People post-stroke also admit to have decreased their level of physical activity because of the restriction measures, fear of infection by Sars-cov2 and their lack of motivation and self-efficacy, which are well documented barriers of physical activity (Ezeugwu et al, 2017; Thilarajah et al, 2018). Studies among the general population and people with a neurological condition also showed, in several countries, a significant reduction of the levels of physical activity (Balci et al, 2021; Varela et al, 2021). Our findings suggest that receiving some sort of guidance from health professionals seemed to be perceived as facilitator to maintain their focus and motivation in the rehabilitation. Moreover, carrying out physical activity alone appeared to decrease their motivation. The importance of involving peers or others (e.g., family, friends, activity with a domestic animal), digitally or in-person, may be an important strategy for health professionals to consider when planning recommendations for people post-stroke.

### Limitations, Application and Future Research

Some limitations might be considered and mitigated in future research. Stroke people with aphasia were not included in our study, which does not allow the findings to be transferred to this subgroup of people post-stroke. Our findings should be considered in terms of transferability to similar contexts in the context in which the study was carried out (Johnson and Waterfield, 2004). Additionally, no respondent validation was performed (participants revision of the data collection and/or analysis), due to time constraints. It would contribute to increase the credibility of the study findings.

## Conclusion

This study provided valuable insights into the perceptions of both people post-stroke and informal caregivers regarding the impact of COVID-19 pandemic on their health/ care. According to the findings of this study, people post-stroke and informal caregivers highlighted the need for support in order to learn new skills, manage challenges and guide decisions. The need for self-management support increased due to the changes in their daily routines, early hospital discharges, cessation of “non-urgent” social and health services and the reduced levels of physical activity. Participants also valued new forms of communication, such as digital solutions that enable remote support strategies. Even prior to the pandemic, in the European Stroke Action Plan 2018-2030, self-management programs were identified as a development priority (Norrving et al, 2018). People post-stroke and their informal caregivers should be trained to the transition between hospital discharge and the return to the community. It is well documented in previous literature that the training of informal caregivers provides competence and reduces their physical and physiological burden (Cameron et al, 2013; Pindus et al, 2018). Telerehabilitation may also be a beneficial approach to the rehabilitation of people post-stroke (Laver et al, 2020; Lee et al, 2021; Markus and Brainin, 2020; Portnoy et al, 2020).

## Data Availability

All data produced in the present study are available upon reasonable request to the authors.

